# Characteristics associated with COVID-19 vaccine uptake among adults in England (08 December – 17 May 2021)

**DOI:** 10.1101/2021.08.27.21262422

**Authors:** Elise Tessier, Yuma Rai, Eleanor Clarke, Anissa Lakhani, Camille Tsang, Ashley Makwana, Heather Heard, Tim Rickeard, Shreya Lakhani, Partho Roy, Michael Edelstein, Mary Ramsay, Jamie Lopez-Bernal, Joanne White, Nick Andrews, Colin Campbell, Julia Stowe

## Abstract

**Objective:** To determine characteristics associated with COVID-19 vaccine coverage among individuals aged 50 years and above in England since the beginning of the programme.

**Design:** Observational cross-sectional study assessed by logistic regression and mean prevalence margins.

**Setting:** COVID-19 vaccinations delivered in England from 08 December 2020 – 17 May 2021.

**Participants:** 30,624,257/ 61,967,781 (49.4%) and 17,360,045/ 61,967,781 (28.1%) individuals in England were recorded as vaccinated in the National Immunisation Management System with a first dose and a second dose of a COVID-19 vaccine, respectively.

**Interventions:** Vaccination status with COVID-19 vaccinations.

**Main Outcome Measures:** Proportion, adjusted odds ratios and mean prevalence margins for individuals not vaccinated with dose 1 among those aged 50-69 years old and dose 1 and 2 among those aged 70 years old and above.

**Results:** Among individuals aged 50 years and above, Black/African/Caribbean ethnic group was the least likely of all ethnic groups to be vaccinated with dose 1 of the COVID-19 vaccine. However, among those aged 70 years and above, the odds of not having dose 2 was 5.53 (95% CI 5.42 to 5.63) and 5.36 (90% CI 5.29 to 5.43) greater among Pakistani and Black/African/Caribbean compared to White British ethnicity, respectively. The odds of not receiving dose 2 was 1.18 (95% CI 1.16 to 1.20) higher among individuals who lived in a care home compared to those who did not. This was the opposite to that observed for dose 1, where the odds of not being vaccinated was significantly higher among those not living in a care home (0.89 (95% CI 0.87 to 0.91)).

**Conclusions:** We found that there are characteristics associated with low COVID-19 vaccine coverage. Inequalities, such as ethnicity are a major contributor to suboptimal coverage and tailored interventions are required to improve coverage and protect the population from SARS-CoV-2.

**Article summary:** *Strengths and Limitations of this study:*

- This is the is the first study assessing characteristics associated with COVID-19 vaccine coverage for all individuals aged 50 years and above in England.
- This study uses data from the National Immunisation Management System (NIMS) which is based on all individuals in England with a registered NHS number.
- This centralised national system captures individual level data for both vaccination status and demographic characteristics and allows for linkage to other datasets such as health care worker and care home resident status.
- This study does not include those without an NHS number and, therefore, it is possible we have underestimated the number of vaccines delivered and odds of not being vaccinated for characteristics such as ethnic groups where we have seen the greatest impact.
- Residual errors in data entry on the point of care apps at the vaccination sites may have also occurred, though these errors are not likely to be widespread.

## Background

The United Kingdom (UK) was the first country in the world to approve a COVID-19 vaccine to be used in response to the pandemic, getting a head start on the roll-out of its COVID-19 vaccination programme ^1^. On December 08 2021, the UK launched its COVID-19 vaccination programme with the aim of reducing COVID-19 mortality and hospitalisations among those at highest risk. There are currently three vaccinations that the Medicines and Healthcare products Regulatory Agency (MHRA) have authorised; the Pfizer/BioNTech vaccine (offered from 08 December 2021), the AstraZeneca (Oxford) vaccine (offered from 04 January 2021) and the Moderna Tx (offered from 13 April 2021) ^2-4^.

To ensure the reduction of mortality from SARS-CoV-2 infection and protect the healthcare system, the Joint Committee on Vaccination and Immunisation (JCVI), an independent expert advisory committee which advises the UK health departments on vaccination, initially recommended extending the interval between doses up to 12 week in order to vaccinate a greater number of people sooner with the first dose of the COVID-19 vaccine ^5 6^. The JCVI also recommended that the vaccination programme be rolled out in phases beginning on 08 December 2021, for those aged 80 years old and above and for frontline healthcare workers ^6^ gradually opening vaccination eligibility. This was followed by phases two to nine in descending age groups along with those who were identified as clinically extremely vulnerable and those classified at risk.

NHS England with support from Public Health England (PHE), the national public health agency, publish COVID-19 vaccination counts and denominators by region, age, ethnicity and for care home residents using a tracker tool ^7^. Previous research on characteristics for low vaccine coverage in England have shown that deprivation and ethnicity are associated with lower coverage ^8-10^. There is little to no data collected on vaccine coverage for individuals living in care homes, though barriers exist in achieving high influenza uptake in care homes such as care home size, geographical location, working relationships with primary care and pharmacies ^11^. These studies and guidance assess routine vaccines delivered in England which are primarily delivered through general practices, pharmacies and schools. With the ongoing COVID-19 pandemic, the rapid development of COVID-19 vaccines and the urgency to rapidly roll out the programme in various settings including mass immunisation sites, it is unknown whether the same characteristics associated with vaccine coverage for routine programmes are associated with low coverage for COVID-19 vaccines.

The aim of this study is twofold:

1. To describe the number of individuals eligible for a COVID-19 vaccine in the first phase of the roll-out that have been vaccinated with a single or two doses of COVID-19 vaccine by age, sex, geographical location, vaccine type, ethnicity, deprivation, urban or rural setting and programme week.
2. To determine whether there are any characteristics independently associated with low vaccine coverage for dose 1 and dose 2 of COVID-19 vaccines.

## Methods

### Data source

A National Immunisations Management System (NIMS) capable of recording any vaccination regardless of point of delivery has been used for the response to the pandemic to collect information about COVID-19 vaccines delivered across England. Individuals who present to a vaccination site, such as general practice, pharmacy, or hospital, provider and receive a COVID-19 vaccine will have their vaccine event information recorded on a Point of Care application. PHE receives data which is linked to demographic data obtained from the NHS (e.g., gender, date of birth), using the individual’s unique NHS number ^12^. PHE uses these data for monitoring vaccine safety, effectiveness and coverage. All variables used for vaccine coverage are described in Supplementary Table 1. Age was calculated for all individuals based on their age on 31 March 2021.

### Patient and Public Involvement

No patients were involved in the design or execution of the study

### Study population

NIMS data was extracted on 17 May 2021 to assess vaccination status of all individuals aged 50 to 69 years old for a first dose of COVID-19 vaccine and of all individuals aged 70 years old and above vaccinated with a first and a second dose of a COVID-19 vaccine. The programme is still being rolled out among younger cohorts therefore not all 50-59 years olds had not been offered a second dose of COVID-19 vaccine at the time of data extraction. Individuals recorded in the NIMS must have an NHS number in order to link the population denominator and vaccination event files. All individuals with a death recorded were excluded from the analyses for the purpose of calculating coverage in the living resident population aged 50 and above on 31 March 2021.

### Vaccine coverage

Vaccine coverage was calculated by dividing the total number of individuals with a recorded NHS number in the dataset who were vaccinated with 1 dose and 2 doses of COVID-19 vaccine since the beginning of the vaccine roll-out on 08 December 2020 (numerator) by the total number of individuals with a recorded NHS number in the dataset (denominator).

### Proportion of unvaccinated individuals

The proportion of individuals not vaccinated for dose 1 and/or not vaccinated with dose 2 was calculated by dividing the total number of individuals unvaccinated or with a single dose of the COVID-19 vaccine since the beginning of the vaccine roll-out on 08 December 2020 (numerator) by the total number of individuals with a recorded NHS number in the dataset (denominator).

### Descriptive analyses

The proportion of individuals vaccinated and unvaccinated was described by programme week, starting at week 1 (beginning on 07 December 2020). Cumulative vaccine uptake has been calculated by programme week. Furthermore, the number of and proportion of individuals unvaccinated were aggregated by age groups, region, rural/urban classification, ethnicity, for individuals clinically extremely vulnerable, for individuals over 65 years old living in a care home, for individuals less than 65 years old and a healthcare worker.

### Statistical analyses

Statistical analyses were conducted for individuals eligible for a COVID-19 vaccine in England in Phase 1, at the beginning of the roll out. The analyses were conducted for those vaccinated with dose 1 and dose 2 for individuals aged 70 years old and above, and for dose 1 among individuals aged 50 to 69 years old.

To assess the odds of not being vaccinated or not being vaccinated with dose 2, univariable logistic regression models for each characteristic were fitted with the binary outcome of vaccination status (not vaccinated/ vaccinated with a single dose and vaccinated with dose 1/ vaccinated with two doses). A multivariable logistic regression model was conducted adjusting for all other characteristics.

The adjusted mean prevalence margins and 95% confidence intervals of being unvaccinated for each characteristic within the multivariable model fit were calculated using the adjusted ratios.

All statistical analyses were conducted using Stata 15.1.

## Results

### Descriptive results

As of 17 May 2021, a total of 30,624,257/ 61,967,781 (49.4%) and 17,360,045/ 61,967,781 (28.1%) individuals were vaccinated with a first dose and a second dose of a COVID-19 vaccine, respectively. The number of individuals vaccinated varied by programme week, plateauing at weeks 11 and 24 for dose 1 and 2 among those aged 70 years old and above, and week 23 for dose 1 among those aged 50 to 69 years old (Figures 1 and 2). Those aged 50 to 69 were still eligible for their second dose and had not yet plateaued at the time the data was extracted.

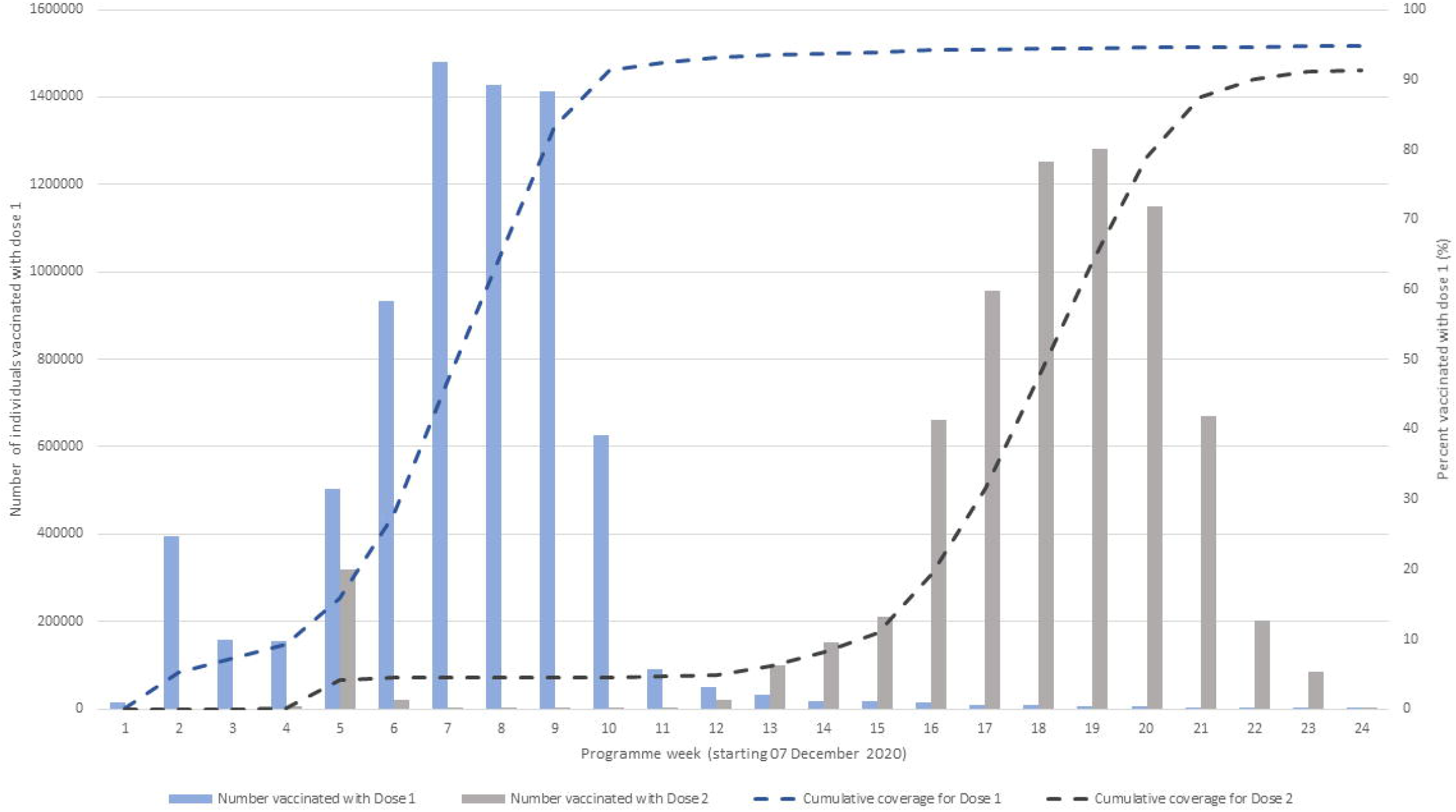

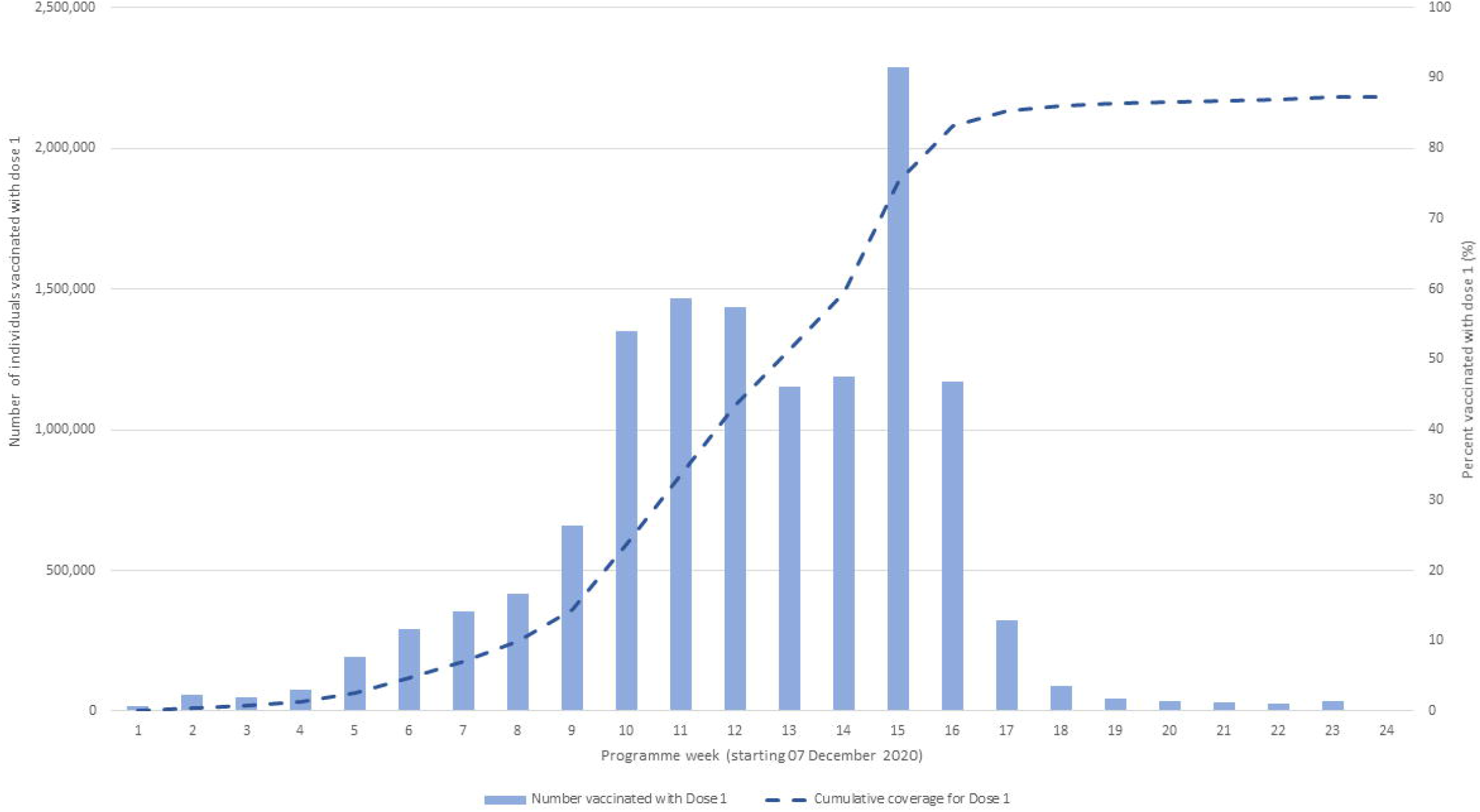

The proportion of individuals unvaccinated varied by age, dose and region; London consistently had the highest proportion of individuals being unvaccinated (Figure 3). Among the total population aged 50 and older, the North East of England had the lowest proportion of unvaccinated individuals (Figure 3).

### 70-year olds and above unvaccinated

Among individuals aged 70 years old and above, all the characteristics were significantly associated with not being vaccinated in both the univariable and multivariable logistic regression analyses.

The odds of not being vaccinated was higher among males and those aged 75-79 and 80 years old and above than the baseline of 70-74 years old (Table 1). The odds of not being vaccinated with 1 or 2 doses was higher in urban areas; particularly in London with an increased odds of 2.30 (2.27 to 2.33) and 1.96 (1.94 to 1.98) odds for not being vaccinated for both dose 1 and dose 2, respectively (Table 1).

**Table 1.**
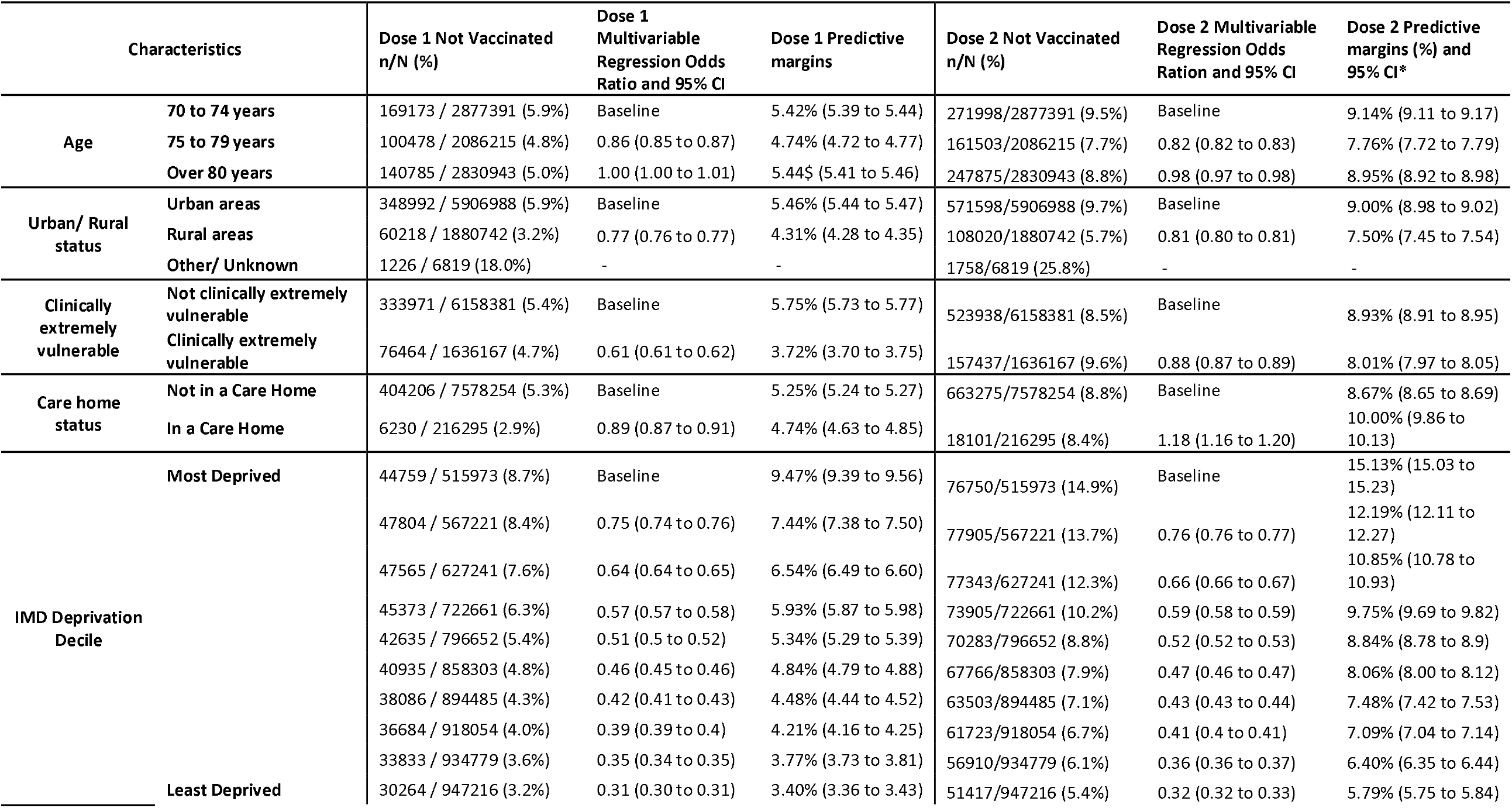
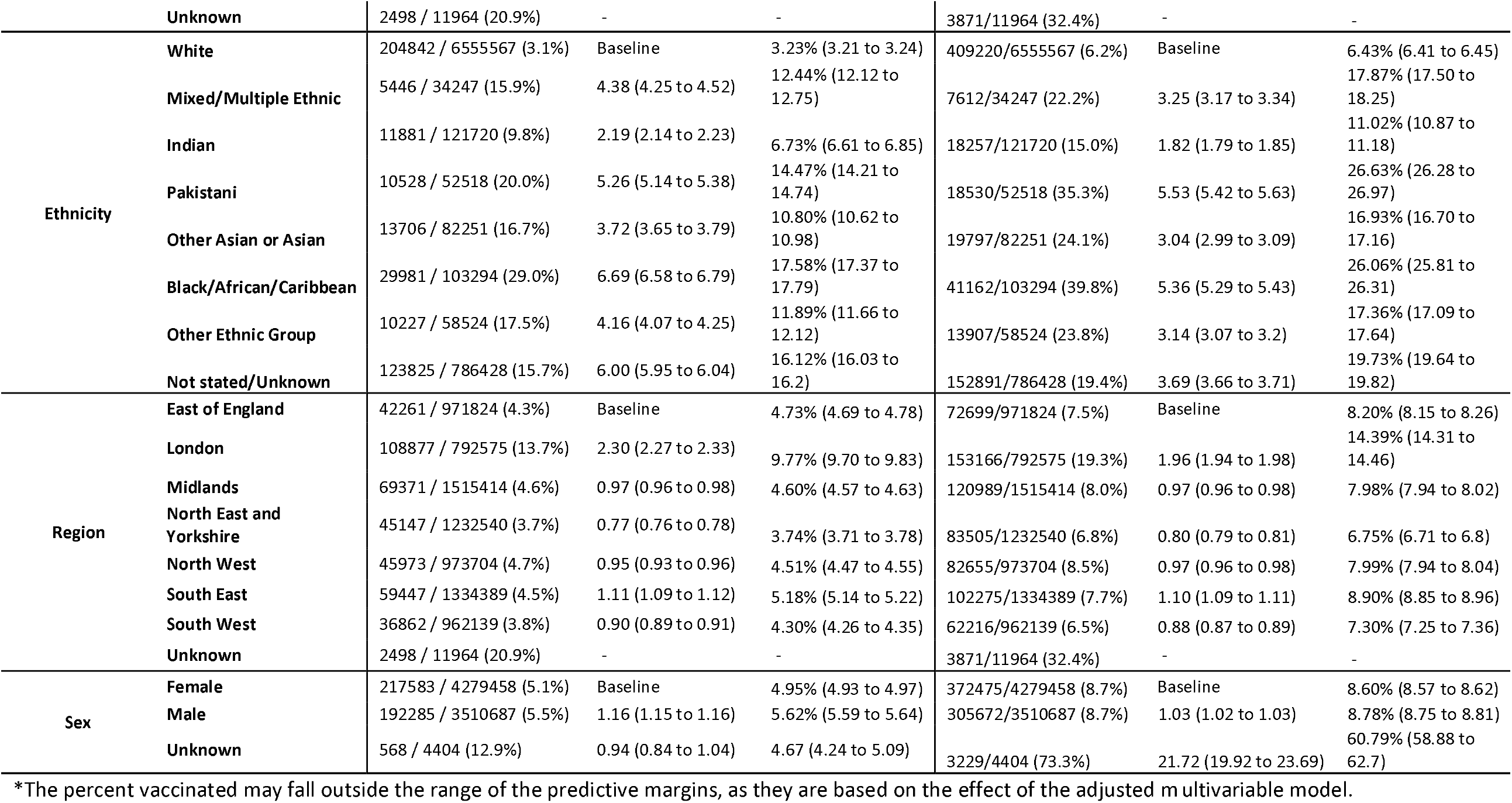
Number and proportion of individuals aged 70 years old and above not vaccinated for dose 1 and dose 2 of the COVID-19 vaccine between 8 December 2020 – 17 May 2021 in England and the odds of not being vaccinated from a multivariable logistic regression and mean adjusted prevalence from the model fit.

Among those clinically extremely vulnerable, the odds of not being vaccinated was reduced (OR 0.61 (95% CI 0.61 to 0.62)) and (OR 0. 88 (95% CI 0.87 to 0.89)) for dose 1 and dose 2, respectively compared to those with no significant underlying health conditions.

The odds of not being vaccinated among those living in a care home was also reduced for dose 1 (OR 0.89 (95% CI 0.87 to 0.91)) compared to those not living in a care home. However, the odds of not being vaccinated for doses 2 of the COVID-19 vaccine was 1.18 (95% CI 1.16 to 1.20) greater. The greatest odds of not being vaccinated among 70 years olds was highest among those living in the most deprived areas and among Black/African/Caribbean ethnicities. Among the Black/African/Caribbean ethnicity, the odds of not having a first dose was 6.69 (95% CI 6.58 to 6.79) greater than White British ethnicity. The odds of not being vaccinated with a second dose was 5.53 (95% CI 5.42 to 5.63) greater among the Pakistani ethnicity, followed by the Black/African/Caribbean ethnicity with an odds of 5.36 (95% CI 5.29 to 5.43).

The mean prevalence of being unvaccinated for each characteristic within the adjusted multivariable model showed increased prevalence among the characteristics with the highest odds of being unvaccinated. The highest prevalence was among the Black/African/Caribbean ethnic group for dose 1 with a mean prevalence of 17.6% unvaccinated. The prevalence among those not being vaccinated with dose 1 was 26.6% and 26.1% among the Pakistani and Black/African/ Caribbean ethnicities, though the 95% confidence intervals do overlap between the two ethnicities (Table 1).

### 50-69-year olds unvaccinated

Of those aged 50-69 years old, all the characteristics were significantly associated with not being vaccinated in both the univariable and multivariable logistic regression analyses.

Similarly to those aged 70 and above, the odds of not being vaccinated was higher among males and increased in the younger populations (Table 1). The odds of not being vaccinated was higher in urban areas and particularly in London where there was an increased odds of 1.85 (95% CI 1.84 to 1.86) (Table 2).

**Table 2.**
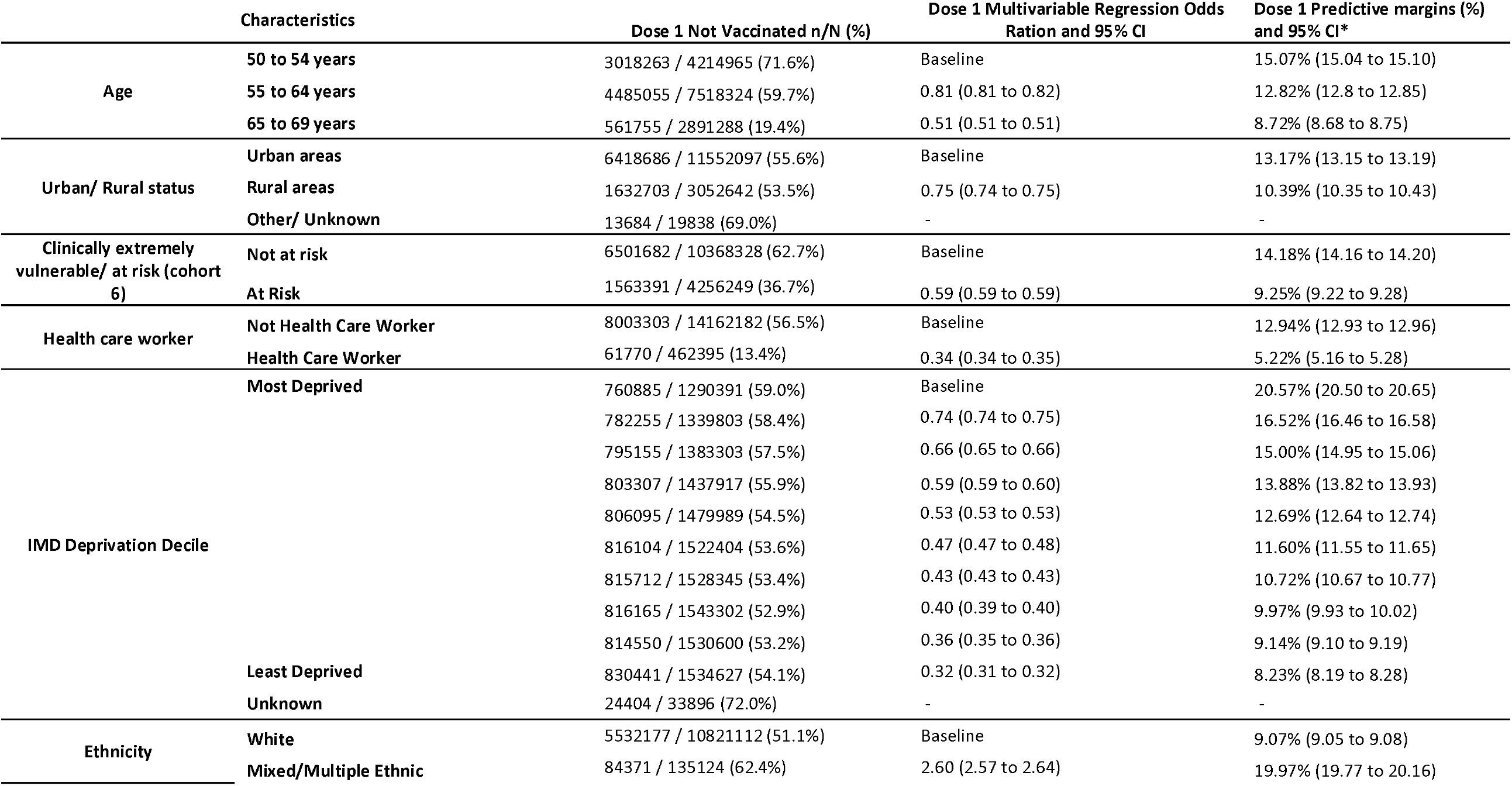
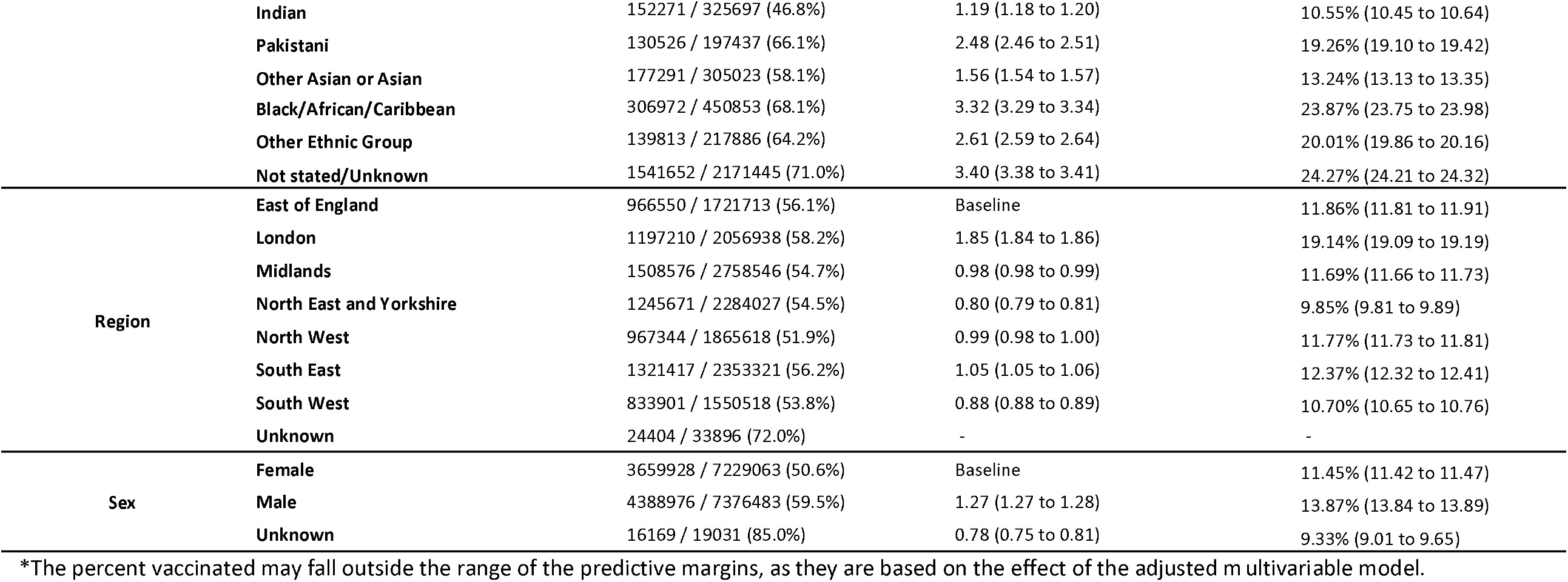
Number and proportion of individuals aged 50-69 years old not vaccinated for dose 1 of the COVID-19 vaccine between 8 December 2020 – 17 May 2021 in England and the odds of not being vaccinated from a multivariable logistic regression and mean adjusted prevalence from the model fit.

The greatest odds of not being vaccinated with dose 1 among 50-69 year olds was highest among those with an unknown or not stated ethnicity and Black/African/Caribbean ethnicity which had a 3.40 (95% CI 3.38 to 3.41) and 3.32 (95% CI 3.29 to 3.34) increased odds of being unvaccinated compared those of White British ethnicity (Table 2).

Both the clinically extremely vulnerable/ at risk and healthcare workers had decreased odds of not being vaccinated than those that were not in these groups (OR 0.59 (95% CI 0.59 to 0.59) and 0.34 (95% CI 0.34 to 0.35), respectively) (Table 2), showing a protective effect among those at highest risk of SARS-CoV-2 disease.

The highest predictive margin for not being vaccinated after adjusting for all variables in the multivariable logistic regression model was among the Unknown/Not Stated and Black/African/Caribbean ethnic groups with a prevalence of 24.7% and 23.9% unvaccinated, respectively (Table 2).

## Discussion

As of 17 May 2021, 49.4% of the population aged 50 and above in England had received a first dose of the COVID-19 vaccine and 28.1% had their second dose. Vaccine coverage for dose 1 among those aged 50 years old and for dose 2 for those aged 70 years old and above have plateaued. A significant increase in the number of individuals aged 50-69 years old occurred in week 15 of the programme (the week commencing the 15 March 2021), when those aged 50 – 69 became eligible for the vaccine ^13 14^. Individuals aged 50 to 69 vaccinated in earlier weeks were most likely to have been healthcare workers, individuals clinically extremely vulnerable/Cohort 6 at risk for severe coronavirus disease ^15^.

The overall proportion of individuals eligible for the vaccine but not vaccinated was highest in London, followed by areas in the Midlands and East of England. These results coincide with findings from studies assessing influenza and shingles vaccination programmes ^8 9^, which offered vaccines to individuals in similar populations such as elderly individuals and those at risk, where lower coverage in London and in urban areas is observed.

Results from the multivariable logistic regression model and the predictive margins for two doses among those aged 70 years old and above and for one dose among those aged 50-69 indicate an increased odds and predicted margins of being unvaccinated with lower age groups. Should lower coverage among younger adults continue to be observed as the programme continues to roll-out, efforts will need to be made to address this. Moreover, after adjusting for all variables in the multivariable models the odds of being unvaccinated was higher among males compared to females, in urban areas compared to rural areas, and highest in London compared to all other regions in England among all individuals aged 50 years old and above. An overall reduced odds of not being vaccinated was observed among those clinically extremely vulnerable/ at risk aged 50 years and above and among healthcare workers aged 50 to 69 years old. These individuals were all eligible for the vaccine early within the programme.

Individuals aged 70 years old and above living in a care home had a reduced odds of not being vaccinated for dose 1 compared to those not living in a care home. In contrary, care home residents had an increased odds of not being vaccinated for the second dose compared to those living in a care home. The delivery of COVID-19 vaccines to care homes was primarily carried out by mobile teams which required a lot of logistics and coordination ^16^. The delivery of second doses of COVID-19 vaccinations in care homes may be lower due to following up individuals who were not vaccinated or partially vaccinated and moved into a care home after or between mobile vaccination unit visits. It is important to further investigate the increased odds of not being vaccinated with dose 2 in care homes, as residents have been disproportionately affected by SARS-CoV-2 ^17^. It is unlikely that the death rates are associated with low coverage as deaths are recorded in a timely manner in the NIMS ^18^, though it is possible that care home residents might have had COVID-19 or another illness at the time of offer, thus causing a lag in the number of second doses received.

Both the highest odds and the predictive margins for not being vaccinated for dose 1 among all individuals in the study and for dose 2 among those aged 70 years old and above were observed in the most deprived and among the Black/African/Caribbean ethnicity. This coincides with findings assessing COVID-19 vaccine coverage for those aged 70 years old and above using the 2011 ONS denominator population estimates ^19^ and using general practice records for population estimates based on registrations for two of the three GP IT System Suppliers in England ^20^. Furthermore, among those aged 70 years old and above, the odds and prevalence of being unvaccinated among those of Pakistani ethnicity further increased for dose 2. Our findings highlight the structural and complex interplay of ethnicity and deprivation which has also been observed with finding on SARS-CoV-2 infection, hospitalisations and mortality ^21 22^.

### Strengths and Limitations of this study

Our study has several strengths; this is the is the first study assessing characteristics associated with COVID-19 vaccine coverage for all individuals aged 50 years and above in England and one of the first studies globally assessing COVID-19 vaccine coverage. Our study also uses data from the NIMS which is based on all individuals in England with a registered NHS number which is likely to be more complete than other datasets used to estimate COVID-19 vaccine coverage. Furthermore, immunisation registers have been proven to be fundamental when assessing and protecting the population, can be used for linkage to health-outcome databases and can play a key role in the delivery of a national immunisation programme ^23-25^. This is the first time England has developed a centralised national system capturing individual level data for both vaccination status and demographic characteristics. Previous studies assessing factors influencing vaccine coverage in England have been based on aggregate general practice-level data where estimates such as deprivation were based on the general practice post code. Having individual level data such as for frontline healthcare workers and care home residents allowed us to link individual NHS numbers to properly account for these individuals which is not available in similar studies or in general practice records.

We are unable to capture details on the total number of individuals without an NHS number and, of those who had not received a vaccine, therefore, it is possible we have underestimated the number of vaccines delivered and odds of not being vaccinated for characteristics such as ethnic groups where we have seen the greatest impact. The proportion of individuals aged 50 years and above with no NHS number is expected to be marginal. Furthermore, it is possible that there could be residual errors in data entry on the point of care apps at the vaccination sites, though these errors are not widespread. Though the NIMS was rapidly set up for monitoring COVID-19 vaccinations, it was piloted with influenza vaccinations delivered in the 2020/21 influenza season and the trends observed in our study align with other immunisation programmes.

## Conclusions

This study provides evidence that in England, being male, being in a younger age group, belonging to certain minority groups, living in urban setting or being a care home resident were associated with low COVID-19 vaccine coverage. The largest odds of not being vaccinated was observed among those of Black/African/Caribbean ethnicity and those in the most deprived decile. It is of utmost importance to reduce inequalities in vaccine coverage, particularly among Black, Asian and Minority Ethnic groups and care home residents who have been most impacted by the SARS-CoV-2 infection. As vaccine coverage increases in England, tailored strategies taking into consideration barriers specific to these under-vaccinated groups should be designed and implemented in order to improve coverage.

## Supporting information

Ethics letter

## Data Availability

No additional data available.

## Acknowledgements

We would like to thank the Public Health England COVID-19 Data Science and ImmForm Teams, NHS England, NHS Digital and System C for their roles in developing and managing vaccination systems and datasets.

## Contributor and guarantor information

Elise Tessier is the guarantor of the paper and confirms that the manuscript is honest, accurate and transparent. There are no important aspects of the paper omitted.

## Ethics Approval

Surveillance of covid-19 vaccination data is undertaken under Regulation 3 of The Health Service (Control of Patient Information) Regulations 2002 to collect confidential patient information (www.legislation.gov.uk/uksi/2002/1438/regulation/3/made) under Sections 3(i) (a) to (c), 3(i)(d) (i) and (ii) and 3(3).

## Data Sharing Statement

No additional data available.

