## Supplementary Table 1 for "Characteristics associated with COVID-19 vaccine uptake among adults in England (08 December – 17 May 2021)"

**Supplementary Table 1.** Data fields provided by NIMS and other data sources used for vaccine coverage PHE for Vaccine Coverage

| **Data source** | **Data Field** | **Field explanations** |
| --- | --- | --- |
| **Population denominator data file** | First name | Limited to 50 characters |
|  | Surname | Limited to 50 characters |
|  | Date of birth | DD/MM/YYYY |
|  | NHS number | 10 Digit number without space |
|  | Sex | Male; Female; Unknown |
|  | Ethnicity | Based on the 2001 ethnic category codes: ETHNIC CATEGORY CODE 2001 (datadictionary.nhs.uk) |
|  | Postcode | Post code with no spaces |
|  | General practice code | Code of the individuals GP practice |
|  | Flag for individuals clinically extremely vulnerable | Based on the English Shielded Patient List |
|  | Flag for frontline healthcare and social care workers | Provided by NHS Business Services Authority for all staff who are directly employed by the NHS organisations using the Electronic Staff Record (ESR). |
|  | Clinically Extremely Vulnerable | Provided by NHS Digital and is a record of vulnerable patients thought to be at high risk of complications from COVID-19. The data heavily relies on data linkage using the NHS number to extract data from the GP electronic health record (EHR), Hospital Episode Statistics (HES), and the QCOVID risk stratification assessment. Specific rule logic can be found here: https://digital.nhs.uk/coronavirus/shielded-patient-list/methodology/rule-logic |
|  | Flag for individuals 16-65 at risk (Cohort 6) | Provided by NHS Digital and is based on a list of NHS numbers extracted from the EHR based on the national PHE PRIMIS SNOMED specification. This includes all those who are in a clinical risk group and coded as a carer within the EHR. For more information on the national specification can be found here: https://www.nottingham.ac.uk/primis/covid-19/covid-19.aspx |
| **Vaccination events data file** | NHS Number | 10 Digit number without space |
|  | Date of vaccination administration | DD/MM/YYYY |
|  | Location Code | The unique code for the location where the vaccination event occurred |
|  | Location Name | The name of the location where the vaccination event occurred |
|  | Vaccine code | SNOMED CT concept code for the Vaccine Code |
|  | Vaccine Procedure Code | SNOMED CT concept code for the Vaccination Procedure |
|  | Route of vaccination | SNONED CT concept code for the route the vaccine was administered. |
|  | Body Site | SNOMED CT concept code for the for the body site where the vaccination was administered |
|  | Batch number | Vaccination’s batch number (from the physical product) |
|  | Manufacturer | Vaccination manufacturer name |
| **Externally provided data sources** | Index of Multiple Deprivation | From 2011 Census - linked by individuals post code. |
|  | Care home status | Unique Property Reference Numbers and NHS-Addresses are used to link to the care home Care Quality Commission addresses. These are the used to linked to the Master Patient Index provided by NHS England and Improvement. The list of individuals in a care home is updated monthly. |
| **Derived variables for coverage** | Age as of 31 March 2021 | Calculated based on DOB |
|  | Manufacturer | Manufacturer is allocated using the first few characters of the Batch Number fields once any leading spaces and prefixes of Batch or BN are removed from the text string. Where it is not possible to identify the manufacturer using the first few characters of the trimmed Batch Number string, records where the batch number includes the string ‘Pfizer’ are allocated a Pfizer manufacturer code Pfizer, records including the strings ‘AstraZeneca’ (or ‘Astra’ and ‘Zeneca’) are allocated an AstraZeneca Code and records including the strings ‘Moderna’ are allocated a Moderna code.  Finally, where manufacturer cannot be allocated using the above rules the manufacturer specific SNOMED codes are used to allocate manufacturer |
|  | Dose number | Dose 1 should be on or after 08 December 2021. A completed course considered valid if dose 2 is a minimum of 20 days after dose 1 |
|  | Clinically Extremely Vulnerable/Cohort 6 | This is a merged flag for those ages 16-65 that have been coded as at risk, anyone ages 16-69 clinically extremely vulnerable, or categorised as both. |
