## Supplementary material for "Characteristics associated with COVID-19 vaccine uptake among adults in England (08 December – 17 May 2021)": Ethics letter

Public Health England

PHE Research Support and Governance Office
Wellington House
London
SE1 8UG

27th August 2021

www.phe.gov.uk

For the attention of the medRxiv team
Your reference: MEDRXIV/2021/262422

Dear Sir/Madam

**Re: Characteristics associated with COVID-19 vaccine uptake among adults in England (08 December to 17 May 2021)**

This protocol was subject to an internal review in the Research Support and Governance Office and was classified as surveillance that was being undertaken as part of PHE's responsibility to respond to the COVID-19 current pandemic. PHE has legal permission, provided by Regulation 3 of The Health Service (Control of Patient Information) Regulations 2002 to collect confidential patient information (http://www.legislation.gov.uk/uksi/2002/1438/regulation/3/made) under Sections 3(i) (a) to (c), 3(i)(d) (i) and (ii) and 3(3) as part of its outbreak response activities.

As such this work falls outside the remit for ethical review and as no regulatory issues were identified the protocol was approved.

Please let me know if you require any further information.

Yours faithfully

E Coates

Dr Elizabeth Coates
Head of Research Governance
Public Health England
